# ASSESSING THE CASE FOR APPLYING PERSONAL RESPONSIBILITY FOR COVID-19 VACCINATION IN A SOUTH AFRICAN INSURED POPULATION

**DOI:** 10.64898/2026.07.07.26357459

**Authors:** Geetesh Solanki, Francesca Little, Susan Cleary

**Affiliations:** Health Economics Unit, Faculty of Health Sciences, Anzio Road, Observatory, 7925, Cape Town, South Africa; Department of Statistical Sciences, University of Cape Town, South Africa

**Keywords:** COVID-19 vaccination, personal responsibility, luck egalitarianism, health insurance, South Africa, priority-setting

## Abstract

**Background:** Personal choice, the opportunity to select an action from available options, free from external constraint, significantly affects health, risks, and treatment needs. Unhealthy lifestyles contribute substantially to global disease burden, pressuring health systems and reigniting debate about individual responsibility for health. The COVID-19 pandemic brought these debates into sharp focus. In South Africa’s private health sector, vaccine hesitancy persisted where vaccines were freely available, raising questions about fairness when avoidable costs are imposed on others within pooled insurance. This paper develops and applies a structured framework to assess the case for applying personal responsibility(policies linking contributions, coverage, or costs to factors under individual control) using COVID-19 vaccination in a South African insured population.

**Methods:** We employed a multi-part approach drawing on administrative claims and vaccination data from approximately 550,000 insured members (March 2020 to December 2022). We examined vaccination on hospitalisation, utilisation, and expenditure; evaluated fairness from utilitarian (cost-effectiveness and cost-utility) and luck egalitarian (choice vs cost distribution) perspectives; assessed the practical feasibility of responsibility-based mechanisms; and integrated findings through a decision framework.

**Results:** Vaccination was associated with >90% lower hospitalisation risk, shorter stays, and 35 to 55% lower costs. Cost-utility analysis showed vaccination dominated non-vaccination (more QALYs at lower cost). Predictive modelling indicated non-vaccination in higher-risk groups reflected personal choice rather than constrained circumstance. Observed costs exceeded modelled costs (if all vaccinated) by 22%, concentrated among older adults and those with comorbidities. Practical assessment identified a hierarchy from low-risk vaccination rewards to higher-risk surcharges and benefit restrictions.

**Conclusion:** Vaccination was impactful and cost-effective; non-vaccination in higher-risk groups reflected personal choice. Responsibility-sensitive approaches may be justified where choice is demonstrable, impacts clear, and mechanisms proportionate, fair, and feasible. Incentive-based mechanisms offer lower-risk starting points than punitive designs. The framework offers policymakers a tool to weigh accountability, fairness, and solidarity in health-financing policy.

**KEY MESSAGES:**

- A structured assessment framework integrating empirical evidence, ethical reasoning, and practical feasibility can guide judgements on when personal responsibility policies are justified.
- In this South African insured population, COVID-19 vaccination was impactful, cost-effective, and non-vaccination in higher-risk groups reflected personal choice rather than constrained circumstance.
- Responsibility-sensitive approaches are defensible only where choice is demonstrable, benefits are clear, and mechanisms are proportionate, fair, and feasible.
- Incentive-based mechanisms offer lower-risk starting points than punitive or exclusionary designs.

## INTRODUCTION

People make different choices about how to live their lives. Personal choice, understood here as an individual’s opportunity and autonomy to select an action from available options, unconstrained by external parties, encompasses health-related behaviours or decisions that are, or could reasonably be expected to be, within an individual’s control.

Health-related choices significantly affect individuals’ health, the risks they face, and their need for treatment (Cappelen et al., 2006). A large body of evidence shows that unhealthy lifestyles contribute substantially to the high and growing burden of disease globally (2017, Vos et al., 2020, Ferretti, 2015). As a result, scarce medical resources that could be directed at preventing or treating suffering for which no individual is responsible are instead consumed by conditions that could be avoided through changes in individual behaviour(Farrenkopf, 2022).

Personal choice-driven increases in disease burden are placing growing pressure on resource-constrained health care systems globally. This has encouraged theorists and decision makers to ask whether individuals should be required to take greater personal responsibility for their health. In this context, personal responsibility refers to any health policy that links either the relative payment for treatment or the extent of treatment to factors under an individual’s control. The underlying logic is that those who make personal choices that increase their risk of illness or injury should bear a greater share of the associated costs or face limitations on the care they receive (Feiring, 2008, Cappelen et al., 2005).

The question of personal responsibility is particularly relevant in South Africa, where behavioural risk factors contribute substantially to the country’s disease burden (Achoki et al., 2022, Matzopoulos et al., 2022, Bradshaw et al., 2022, Neethling et al., 2022, Groenewald et al., 2022). While South Africa has committed to Universal Health Coverage (UHC) underpinned by principles of access, equity, quality, and financial risk protection, these values may be compromised by the pragmatic measures required to deliver even a modest version of UHC. As the country moves towards National Health Insurance (NHI), it will be critical to establish a clear set of benefits and a transparent framework for selecting them. If quality care cannot be provided for all needs, at what point is it legitimate to shift responsibility from the collective towards individual citizens?

There is a case, therefore, for examining whether personal responsibility should be included as a criterion in benefit package design. However, applying personal responsibility in health is complex and highly contested (Sharkey et al., 2010), requiring careful assessment across multiple dimensions, including impact, fairness, and practicality. While the need for such assessment is clear, the literature offers limited practical guidance on how it should be conducted, and empirical case studies remain rare.

The COVID-19 pandemic reignited longstanding debates about the role of personal responsibility in health systems (Feiring, 2008, Cappelen et al., 2005, Cappelen et al., 2022, Germain, 2020, Cappelen et al., 2006). The global effort to vaccinate against COVID-19 rested on an implicit social contract: individuals were expected to take reasonable steps to protect themselves and others, while society collectively financed and provided the means to do so. Yet in many settings, including South Africa’s private health sector, vaccine hesitancy persisted even where vaccines were freely available(Sewpaul et al., 2023), raising questions about whether it is fair for those who choose not to vaccinate to impose avoidable costs on others within a pooled insurance arrangement.

Personal responsibility in health policy remains deeply contested (Sharkey et al., 2010, Laverty et al., 2018, Minkler, 1999, Traina et al., 2020, Traina et al., 2022). Proponents argue that individuals should bear some consequences for avoidable ill health arising from personal choices, as this promotes fairness and efficient use of limited resources (Minkler, 1999). Critics counter that health-related behaviours often reflect access barriers (including availability, financial risk protection, and acceptability of services) (McIntyre et al., 2009) rather than genuine choice, undermining the fairness of assigning responsibility or imposing penalties (Laverty et al., 2018, Sheeran et al., 2016, Friesen, 2018, Sharkey et al., 2010).

The broader literature reveals significant challenges in using personal responsibility as a prioritisation criterion in health policy (Sharkey et al., 2010, Friesen, 2018). Despite extensive ethical debate, empirical work on how responsibility-sensitive principles could be applied fairly and practically in coverage or benefit design decisions remains limited, particularly in low- and middle-income countries (LMICs) (van der Star et al.). Decision-makers face three options: to introduce personal responsibility while ignoring these challenges; to abandon it altogether as unworkable; or to continue searching for ways to apply it fairly (Sharkey et al., 2010, Buyx, 2008). The first option risks unfairness and social division; the second disregards the growing role of lifestyle and behavioural choices in driving disease burden and resource constraints (2017, Vos et al., 2020, Ferretti, 2015). The third option, based on developing approaches that respond to fairness concerns while recognising the reality of personal choice, offers a potentially constructive path forward (Feiring, 2008, Cappelen et al., 2005).

This study seeks to make progress on the third approach. It develops and applies a structured framework for assessing the case for applying personal responsibility for health, using COVID-19 vaccination within a South African insured population as a case study. COVID-19 vaccination offers a “bounded” test case: it involves a clearly defined preventive action, relatively uniform access across privately insured individuals, and an opportunity to assess both the individual and collective benefits and costs. In this context, choices not to vaccinate can more plausibly be assessed as voluntary or structurally constrained.

Guided by the overarching research question, “Under what conditions, if any, can personal responsibility for health, illustrated through COVID-19 vaccination, be applied fairly in a South African insured population?”, this paper draws together evidence on impact, fairness, efficiency, and practicality from four complementary analyses conducted within the same insured population: a retrospective cohort study of vaccination impact, a cost-effectiveness and cost-utility analysis assessing fairness from a utilitarian perspective, a luck egalitarian assessment of choice and circumstance, and an evaluation of practical policy instruments.. We pursue two aims: first, to propose an assessment framework that integrates ethical reasoning, empirical evidence, and an evaluation of practical feasibility, to guide judgments about when personal responsibility might be justified; and second, to apply this framework to the case of COVID-19 vaccination in a South African insured population.

By combining ethical reasoning with real-world data, this paper offers a structured approach for evaluating when and how responsibility-sensitive policies might be justified without undermining solidarity or access. It aims to contribute to debates on fairness and efficiency in health financing, an issue of growing importance as countries pursue Universal Health Coverage amidst rising disease burden driven by personal choices.

## CONCEPTUAL AND ANALYTICAL FRAMEWORK

### Theoretical basis

Personal responsibility for health is one of the most contested ideas in health policy and bioethics (Minkler, 1999). At its core lies the question of whether individuals should bear consequences for health outcomes that arise, at least partly, from their personal choices. These debates about personal responsibility are situated within broader theories of distributive justice, which address how the benefits and burdens of social cooperation should be distributed. These theories provide the moral basis for decisions about who should receive what level of care, under what conditions, and why.

Libertarianism emphasises individual liberty and minimal state interference, viewing health outcomes as personal matters and resisting redistributive policies (Hanson, 2007, Germain, 2020, Buyx, 2008, Wikler, 2002, Minkler, 1999). Classical egalitarianism instead seeks equality of outcome, contending that fairness requires redressing all inequalities irrespective of cause (Rawls, 2004, Germain, 2020, Cappelen et al., 2005). Communitarianism frames justice in terms of collective values and mutual obligations: individuals are embedded in communities, and fairness reflects shared responsibility for common welfare (Germain, 2019, Buyx, 2008, Germain, 2020).

While all the theories offer distinct perspectives on justice, this research focuses primarily on utilitarianism and luck egalitarianism. Libertarianism, with its focus on minimal state interference, provides an important counterpoint but is inconsistent with the solidarity-based foundations of health insurance and Universal Health Coverage, which depend on redistribution and collective risk pooling. Classical egalitarianism underpins the moral case for equal access but offers little guidance on how to address inequalities arising from personal choice. Communitarianism informs the analysis indirectly, grounding it in concern for solidarity, reciprocity, and collective trust, which act as moral boundaries for any application of personal responsibility.

Within this broader ethical landscape, two complementary perspectives provide arguably the most relevant lenses for assessing personal responsibility in health financing.

Utilitarianism evaluates fairness through consequences, judging policies as morally justified when they produce the greatest good for the greatest number, that is, when they maximise aggregate welfare. In health economics, the utilitarian principle is typically operationalised through cost-effectiveness analysis (CEA) and cost-utility analysis (CUA), which assess whether an intervention represents an efficient use of resources (Edoka et al., 2020). The evaluation used in this study adopts an extra-welfarist approach, focusing on health outcomes measured in natural units (hospitalisations and deaths averted, life-years gained) and quality-adjusted life years (QALYs), rather than on utility in the broader welfare sense (Baltussen et al., Brouwer et al., 2008). While utilitarianism does not directly address individual responsibility, it provides a population-level lens on fairness: a policy that does not produce the greatest good for the population cannot be justified on utilitarian grounds. In the context of this framework, the utilitarian analysis forms one dimension of the fairness assessment, complementing the luck egalitarian focus on individual choice and circumstance.

Luck egalitarianism offers a more direct framework for assessing responsibility. It distinguishes between inequalities arising from factors beyond an individual’s control (brute luck) and those arising from genuine personal choices (option luck) (Cappelen et al., 2005, Baltussen et al., Ekmekçi et al.). According to this view, society has a duty to compensate individuals for disadvantages resulting from brute luck, but inequalities resulting from option luck may be considered acceptable: individuals should bear the consequences of their genuine choices (Albertsen et al., 2014, Segall, 2010). The work of Roemer has been influential in operationalising this distinction, offering a formal framework for separating outcomes due to circumstances from those due to effort.

### Assessment Framework

While normative debates about responsibility are well established, there are few applied frameworks for systematically assessing when and how personal responsibility might be justified in coverage or priority setting decisions. An integrative review (Snyder, 2019) of the literature on priority setting, personal responsibility, and COVID-19 vaccination identified recurring considerations across three domains: (1) the empirical basis for attributing outcomes to individual behaviour (impact); (2) the ethical justification for assigning or withholding responsibility; and (3) the practical and procedural implications and feasibility of doing so within health systems.

Drawing on these findings and the theoretical foundations outlined above, we developed a four-part framework to assess whether personal responsibility for COVID-19 vaccination can be justified empirically, ethically, and practically. The framework, summarised in Table 1, comprises four components addressing impact, fairness from both utilitarian and luck egalitarian perspectives, and practical feasibility. Together, these components provide a structured approach for integrating empirical evidence, ethical reasoning, and practical considerations to assess the overall case for applying personal responsibility.

**Table 1.** Framework for assessing the case for applying personal responsibility for COVID-19 vaccination.

| Framework Component | Key Question | Theoretical Lens/<br>Ethical Focus | Analytical Approach/<br>Data Source | Purpose / Interpretation |
| --- | --- | --- | --- | --- |
| **Part 1: Impact** | Does COVID-19 vaccination reduce hospitalisation, utilisation, expenditure and mortality? | Provides the empirical foundation for the responsibility assessment | Retrospective cohort analysis using medical claims data to estimate effect of vaccination on hospitalisation risk, utilisation, and expenditure | Establishes whether vaccination has a measurable impact — a prerequisite for considering personal responsibility. |
| **Part 2: Fairness (Utilitarian Perspective)** | What is the cost-effectiveness and cost-utility of COVID-19 vaccination? | Does vaccination represent an efficient use of resources and an intervention that leads to increased overall health (extra welfarist approach) (Edoka et al., 2020, Baltussen et al.) | Cost-utility and cost effectiveness analysis using a Markov model to estimate hospitalisations and death averted, LYs and QALYs gained, incremental cost-effectiveness and cost utility ratios for vaccination vs non vaccination | Assesses whether vaccination represents an efficient use of pooled insurance resources and produces the greatest good for the population. Together with the luck egalitarian assessment (Part 3), this forms the fairness component of the overall evaluation. |
| **Part 3: Fairness (Luck Egalitarian Perspective)** | Was non-vaccination due to personal choice or constrained circumstance?<br>How were costs and benefits distributed? | Fairness and equality (luck egalitarianism) | Predictive modelling of vaccination probability to distinguish choice from circumstance; observed vs. predicted (if vaccinated) costs of vaccinated and unvaccinated groups to assess excess cost burdens | Evaluates fairness by distinguishing non-vaccination due to personal choice (option luck) from constrained non-vaccination (brute luck) and by examining distributional equity. |
| **Part 4: Practical considerations** | How might personal responsibility be operationalised in an insured environment?<br>What are the implications? | Procedural and institutional fairness; solidarity and feasibility | Identify potential instruments (e.g., differential premiums, incentives, conditional coverage) in an insured environment based<br>Assess potential impact, ethical implications, implementation hurdles and risk to arrive at overall viability of the instrument<br>Assessment based on finding of this research and published evidence. | Identifies and evaluates feasible mechanisms, their ethical implications, and implementation challenges associated with each. |

The four components are analytically distinct yet interdependent. The impact component establishes the empirical foundation; the fairness components (luck egalitarian and utilitarian) assess ethical defensibility from complementary perspectives; and the practical implications component evaluates whether instruments would be feasible and proportionate in real-world policy settings. Together, these dimensions integrate ethical reasoning with empirical evidence, providing a systematic, transparent approach to assessing responsibility in health financing.

## METHODS AND DATA SOURCES

### Study design

This study synthesises the findings of 4 analyses designed to evaluate the case for applying personal responsibility for COVID-19 vaccination in a South African insured population. Each analysis corresponds to one element of the four-part framework and addresses a specific aspect of the overall question. The impact analysis has been published (Solanki et al., 2025); the economic evaluation and luck egalitarian analyses are under review (Solanki et al., Solanki et al.). A summary of the components is presented below.

### Impact analysis (Part 1)

A retrospective cohort analysis examining the impact of COVID-19 vaccination on hospitalisation risk, health care utilisation, and hospital expenditure was carried out (Solanki et al., 2025). A Cox Proportional Hazards model was used to estimate the impact of vaccination (non-vaccinated, partly vaccinated, fully vaccinated) on COVID-19 hospitalization risk; and zero-inflated negative binomial models were used to estimate the impact of vaccination on hospital utilization and hospital expenditure for COVID-19 infection, with adjustments for age, sex, comorbidities and province of residence (Solanki et al., 2025).

### Fairness – utilitarian perspective (Part 2)

A state-transition Markov cohort model was developed to assess the cost-utility and cost-effectiveness of partial and full vaccination relative to no vaccination from the insurer’s perspective (Solanki et al.). Transition probabilities were derived from the observed cohort data, and outcomes were expressed as hospitalisations and deaths averted, life-years and QALYs gained, and incremental cost per QALY gained. All costs were estimated in 2022 USD and discounted at 3%. This analysis forms the utilitarian component of the fairness assessment: it evaluates whether vaccination produced the greatest good for the population, that is, whether it represented an efficient use of pooled resources. If vaccination were not cost-effective, it could not be justified on utilitarian grounds.

### Fairness- luck egalitarian perspective (Part 3)

The analysis first examined the extent to which external structural barriers contributed to non-vaccination. This involved a contextual review of policy documents, scheme communications, and regulatory requirements governing vaccine access within the insured population (Solanki et al.). This was accompanied by a two-part empirical analysis. The first part aimed to distinguish between members whose non-vaccination was likely due to personal choice (option luck) and those whose non-vaccination was likely constrained by circumstance (brute luck). Logistic regression models were used to predict each member’s probability of being vaccinated based on demographic and clinical characteristics. Observed and predicted vaccination rates were then compared across probability strata to infer whether non-vaccination reflected personal choice or constrained opportunity. The second part of the analysis estimated the additional financial costs associated with non-vaccination. Cost and utilisation outcomes were modelled using a zero-inflated negative binomial approach to account for the high proportion of members with zero hospitalisation costs. The estimated coefficients were then used to predict expenditure under a counterfactual scenario in which all members were vaccinated. The difference between observed and predicted costs provided an estimate of the excess costs attributable to non-vaccination.

### Practical assessment (Part 4)

The practical assessment aimed to extend the analysis from ethical justification to the identification and evaluation of potential policy instruments through which a health insurer could operationalise personal responsibility for COVID-19 vaccination. Four instruments were assessed, each representing a different point on a coerciveness continuum: vaccination rewards, contribution surcharges, benefit modification, and exclusion of cover. Each instrument was evaluated against three criteria: potential impact on vaccination behaviour and on the distribution of costs, ethical defensibility, and implementation feasibility.

### Overall Assessment

The findings from the four component studies were integrated in three steps: (1) results from each study were mapped onto the corresponding component of the assessment framework; (2) the evidence for each domain was synthesised to assess whether the empirical, ethical, and practical criteria for applying personal responsibility were met; and (3) a decision framework, presented as a policy decision tree, was developed to guide interpretation and application of the evidence (Figure 1). The decision tree operationalises the four-part framework in a structured sequence. It links empirical evidence on vaccination impact, economic efficiency, the distinction between personal choice and constrained circumstance and the resulting fairness of cost distributions, and practical feasibility. It enables decision-makers to determine whether, and under what conditions, personal responsibility for vaccination can be ethically and practically justified.

**Figure 1.**
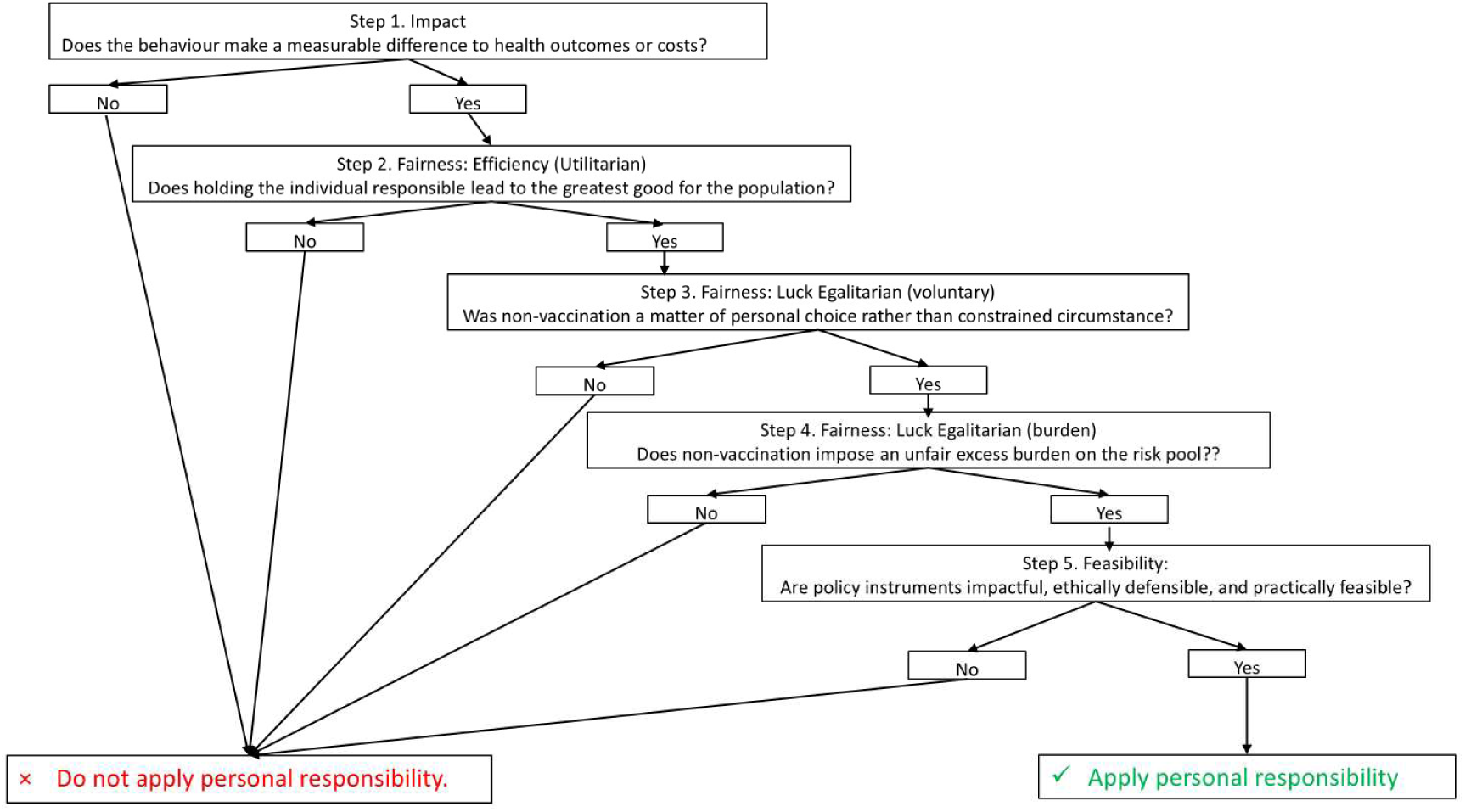
Decision tree deciding on whether to apply personal responsibility for COVID-19 Vaccination.

### Data sources

The empirical analyses drew on de-identified, routinely collected data from a large South African private health-insurance administrator. The dataset comprised approximately 550,000 individuals enrolled between March 2020 and December 2022. It included vaccination records, linked medical and hospital claims, diagnostic codes, and expenditure data, allowing longitudinal assessment of both health outcomes and costs. Use of real-world administrative data permitted estimation of both health-system impact and cost-utility. Because the data reflected observed utilisation and expenditure, findings were directly relevant to policy decisions within private insurance environments.

### Ethical considerations

The data for the studies on which a large part of this evaluation is based were accessed under the terms and conditions set out in the agreement between NMG and the client insurance funds and between NMG and the researcher. The terms set out in these agreements ensured that the access and use of the data was in full compliance with the requirements of the Protection of Personal Information (PoPI) Act in South Africa. Ethics approval for the use of the database to carry out this study was granted by the Human Research Ethics Committee of the Faculty of Health Sciences, University of Cape Town (reference number 606/2023).

## RESULTS

### Overview

The results are organised according to the four components of the assessment framework, followed by an integrated synthesis using the decision framework (Figure 1). Together, these components provide a structured assessment of the empirical, ethical, and practical dimensions of applying personal responsibility for COVID-19 vaccination within a South African insured population as summarised in Table 2.

**Table 2.** Summary of findings of component studies and overall assessment.

| Assessment Dimension | Key Research Question | Summary of Findings | Implications for Personal Responsibility |
| --- | --- | --- | --- |
| **1. Impact** | Does COVID-19 vaccination reduce hospitalisation, utilisation, expenditure, and mortality? | Hospitalisation risk 93–94 % lower for vaccinated members.<br>Hospital utilisation 17–20 % lower.<br>Hospital expenditure 35–55 % lower. | Strong justification.<br>Vaccination provides a clear, quantifiable basis for risk differentiation and creates a financial rationale for considering responsibility-linked policy instruments. |
| **2. Fairness: Utilitarian Perspective** | Does vaccination represent an efficient use of pooled insurance resources — that is, does it produce the greatest good for the population? | Vaccination dominated non-vaccination, improving outcomes at lower cost.<br>ICURs = – \$651 to – \$145 per QALY gained, well below indicative cost-utility thresholds.<br>Probabilistic sensitivity analyses confirmed near-universal cost-saving results. | Strong justification.<br>Vaccination produced the greatest good for the population, maximising collective health and representing an efficient use of pooled insurance resources. |
| **3. Fairness: Luck Egalitarian Perspective** | Was non-vaccination due to personal choice or constrained circumstance, and how were costs and benefits distributed? | Vaccines were free and widely available, and extensive communication campaigns addressed information-related barriers; financial and physical access barriers were substantially reduced.<br>Comparison of observed vs predicted uptake identified groups likely unvaccinated by personal choice.<br>Unvaccinated members incurred 22 % higher costs than predicted under universal vaccination; excess costs concentrated among older adults and those with comorbidities.<br>Among younger, low-risk members, vaccination costs slightly exceeded savings. | Conditional justification.<br>Individuals who were likely to be unvaccinated by personal choice rather than circumstance can be identified.<br>Holding individuals responsible is ethically more plausible where non-vaccination reflects personal choice.<br>The identified cross-subsidy provides a clear focus for corrective or incentive-based policy instruments. |
| **4. Practical considerations** | What policy instruments could operationalise personal responsibility, and what are their practical, ethical, and implementation implications? | Incentive-based rewards – low impact, low risk.<br>Contribution surcharges – higher impact but higher risk.<br>Benefit modification or exclusion – highest impact highest risk strategy | Pragmatic pathway: Scope for action exists, but implementation must be proportionate and incremental, starting with low-risk incentives and supportive approaches.<br>Empirical justification is tempered by ethical and operational constraints. |
| **5. Consolidated Implication** | What is the overall evidence base for considering personal responsibility? | Evidence demonstrates that:<br>Vaccination is impactful,<br>Vaccination represented the most efficient use of collective funds.<br>Non-vaccination due to choice versus circumstance can be systematically assessed<br>Higher costs due to non-vaccination can be quantified<br>Practically feasible if the approach adopted is proportionate and incremental starting with low-risk incentive-driven approaches. | Overall conclusion: A conditional, proportionate case can be made applying personal responsibility, starting with incentive-based, solidarity-consistent mechanisms.<br>The strength of the case is dependent on which ethical perspective(s) is valued. |

### Part 1 – Impact: Does vaccination make a difference?

After adjusting for age, sex, comorbidities, and other confounding factors, vaccination had a marked effect on COVID-19-related illness, hospital utilisation, and costs. Vaccinated members had a 93–94% lower risk of hospitalisation, 17–20% fewer hospital days, and 35–55% lower hospital expenditure compared with those unvaccinated (Solanki et al., 2025). The findings provide clear empirical evidence that vaccination substantially reduces both individual risk and collective system costs. They establish a strong foundation for any consideration of personal responsibility, showing that vaccination status is a behavioural factor that materially influences outcomes. Without such evidence of impact, ethical or practical arguments for responsibility would have no empirical basis (Solanki et al., 2025).

### Part 2 – Fairness: Utilitarian assessment

From a utilitarian perspective, vaccination produced the greatest good for the population, delivering substantial health gains by averting hospitalisations and deaths, increasing life-years and QALYs, and reducing overall expenditure (Solanki et al.). The cost-utility analysis showed that vaccination dominated non-vaccination, being both less costly and more effective. Incremental cost-utility ratios ranged from –$651.28 (partial vaccination) to –$144.73(full vaccination) per QALY gained. Negative ICURs indicate that vaccination produced more QALYs at lower cost than non-vaccination, that is, it was both more effective and cost-saving. Sensitivity analyses confirmed these results, with almost all iterations falling within the cost-saving quadrants, reinforcing the robustness of the findings (Solanki et al.).

### Part 3 – Fairness: Luck egalitarian assessment

Two main findings emerged from the assessment of fairness from a luck egalitarian perspective (Solanki et al.). First, non-vaccination in this insured population was unlikely to result from financial or structural barriers, given that vaccines were free at the point of delivery, widely available, and actively promoted through public and insurer-level communication. Predictive modelling, validated by the close alignment between predicted and observed vaccination rates, suggests that non-vaccination among members with a high predicted probability of vaccination most likely reflected personal choice rather than constrained circumstance. Second, unvaccinated members generated measurable excess health care costs relative to modelled predictions. Observed costs exceeded predicted costs by an average of 22%, with the excess concentrated among individuals with predicted vaccination probabilities above 50%, those for whom non-vaccination is most plausibly due to personal choice. Among younger, low-risk members, however, predicted costs (if vaccinated) slightly exceeded observed spending, as the fixed cost of vaccination was not fully offset by avoided hospitalisation.

### Part 4 – Practical assessment

The practical assessment evaluated four potential policy instruments: vaccination rewards, contribution surcharges, benefit modification, and exclusion of cover against criteria of potential impact, ethical defensibility, and implementation feasibility (Sharkey et al., 2010, Friesen, 2018, Schnaudt et al., 2021).

Vaccination rewards offer moderate impact on vaccination behaviour, with evidence showing some uptake increases in specific contexts (Campos-Mercade et al., 2021, Shen et al., 2024). They are ethically uncontroversial, avoiding charges of harshness or stigma (Sharkey et al., 2010), and implementation is straightforward with low regulatory scrutiny. Overall, rewards are a low-risk, low-impact strategy that does not directly address the identified cross-subsidy.

Contribution surcharges have potentially high impact, with real-world evidence of immediate behavioural response (Fayaz-Farkhad et al., 2024) and direct alignment with individual risk. They raise ethical concerns about harshness and inequity (Sharkey et al., 2010, Friesen, 2018), and may face legal and political risks. Overall, surcharges are a higher-risk, higher-impact strategy.

Benefit modification directly reduces cross-subsidisation, supported by evidence from Part 3 of 127.6% excess costs in high-probability/high-risk groups. However, it raises ethical concerns (Sharkey et al., 2010), may compromise the provider-patient relationship, and conflicts with the South African Prescribed Minimum Benefit regulatory framework which requires medical schemes to cover COVID-19 treatment in full, without co-payments or benefit caps, regardless of vaccination status. Overall, it is a higher-risk, higher-impact strategy that could be legally and ethically challenging.

Exclusion of cover is the highest-risk, highest-impact strategy, theoretically coherent but practically and legally challenging strategy. While Singapore’s approach demonstrates administrative feasibility(Koh et al., 2024), it is likely to raise ethical objections [8, 28] and is in contravention of South Africa’s Medical Schemes Act and Prescribed Minimum Benefit framework which requires medical schemes to provide full cover for COVID-19 treatment to all members regardless of vaccination status.

In conclusion, the empirical case for considering personal responsibility is strong, but the path to implementation is not straightforward and a single, punitive instrument may not be feasible. A phased, ‘middle way’ approach may represent the most feasible way to proceed. Within such an approach, the first step would be the implementation of some form of positive incentives for vaccinating accompanied by communication of the research findings to members, promoting a shared understanding of the collective financial impact of personal choices. If the softer approach of incentivisation and communication fails to mitigate the inequities of the cross-subsidy, the scheme may then consider the more punitive measures. This graduated strategy offers a pathway for applying personal responsibility in a manner that is evidence-based, ethical, and operationally sustainable.

### Part 5. Overall Assessment and Decision Framework

The integrated analysis brings together the empirical, ethical, and practical findings to assess whether personal responsibility for COVID-19 vaccination can be justified within a South African insured population. Using the decision framework as a synthesis tool, the findings point to four key insights regarding the application of personal responsibility for COVID-19 vaccination in this insured population.

- First, the empirical prerequisite for considering any responsibility-based policy was met: vaccination had substantial impact. Vaccination significantly reduced hospitalisation, utilisation, and expenditure (Solanki et al., 2025). Without clear evidence of impact, there would be no empirical basis for considering responsibility, regardless of ethical or practical arguments.
- Second, fairness considerations, assessed from both utilitarian and luck egalitarian perspectives, provided conditional support. From a utilitarian perspective, vaccination produced the greatest good for the population: it dominated non-vaccination on cost-utility analysis, delivering more QALYs at lower cost(Solanki et al.). From a luck egalitarian perspective, non-vaccination in higher-risk groups largely reflected personal choice rather than constrained circumstance, and unvaccinated members incurred 22% higher costs than would have been expected under universal vaccination (Solanki et al.). However, fairness requires that responses be proportionate, recognising the distinction between choice and circumstance (Cappelen et al., 2005, Sharkey et al., 2010).
- Third, feasibility shapes what form responsibility can take. The practical assessment showed that incentive-based and supportive approaches are both operationally feasible and ethically defensible, while punitive mechanisms risk inequity, adverse selection, and loss of trust. A clear hierarchy emerges with vaccination rewards likely to be a low-risk, low-impact strategy; contribution surcharges represent a higher-risk, higher-impact strategy; benefit modification and exclusion of cover are likely to be highest-risk, highest-impact.
- Fourth, taken together, the analysis indicates a qualified, context-dependent case for applying personal responsibility, with enabling incentives as the most viable initial pathway. The conclusions drawn from the framework are inherently value-dependent: the same evidence supports different policy responses depending on whether decision-makers prioritise efficiency, equity, or solidarity. The following discussion situates these results within broader debates on fairness, efficiency, and justice in health-system design and considers their implications for insurers, policymakers, and Universal Health Coverage in South Africa.

## DISCUSSION

This study assessed the overall case for applying personal responsibility for COVID-19 vaccination in a South African insured population. Using a structured, multi-component assessment framework that integrated empirical evidence with ethical reasoning, it examined the impact of vaccination (Solanki et al., 2025), fairness from utilitarian (Solanki et al.) and luck egalitarian (Solanki et al.) perspectives, and the practical feasibility of responsibility-sensitive policy instruments. An assessment framework was developed and applied to move beyond abstract ethical debate and provide a transparent, evidence-based approach for determining when personal responsibility may be ethically justified and operationally feasible.

The findings suggest that applying personal responsibility for COVID-19 vaccination can be ethically and empirically defensible under specific conditions, namely, where vaccination demonstrably improves outcomes, non-vaccination largely reflects personal choice, and policy instruments are proportionate, feasible, and procedurally fair. However, justification weakens when equity and solidarity are prioritised over efficiency or when instruments risk exclusion or inequity.

The analysis highlights the tension between fairness and efficiency in responsibility-sensitive policy raised elsewhere (Norheim, 2016). From a health maximisation perspective, the case for vaccination is unambiguous: it produces the greatest good for the population by preventing avoidable hospitalisations and deaths, reducing costs (Solanki et al., 2025), and maximises collective health (Solanki et al.). From a luck egalitarian standpoint, fairness depends not only on whether non-vaccination reflected choice rather than circumstance, but also on the proportionality of the response. The findings indicate that it is possible to identify individuals for whom non-vaccination reflected personal choice, suggesting that differential incentives may be ethically justified (Solanki et al., Norheim et al., 2009). Yet fairness extends beyond the personal choice/circumstance distinction. Responsibility must remain proportionate to the behaviour’s impact and consistent with solidarity. Modest incentive-based or contributory differentiation may therefore be fair and acceptable, while punitive or exclusionary measures risk undermining equal respect, autonomy, and trust. Responsibility-sensitive policy should thus balance accountability with inclusion, starting with pathways that reward responsible behaviour without punishment or stigma (Daniels et al., Wikler, 2002).

The decision framework highlights that the legitimacy of applying personal responsibility depends on how ethical and practical priorities are balanced. When fairness, efficiency, and feasibility are weighted equally, the case for incentive-based mechanisms is strong. Under efficiency-dominant assumptions, justification strengthens further as vaccination improves outcomes and saves costs. Conversely, when solidarity and equality dominate, justification weakens because the potential social costs of inequity could outweigh efficiency gains. Ultimately, the conclusions reached through this framework are inherently value-dependent: the same empirical evidence can yield different policy judgments depending on whether decision-makers prioritise efficiency, fairness, or solidarity. Those who place greater weight on efficiency may view responsibility-sensitive approaches as justified because they optimise outcomes and resource use, whereas those emphasising fairness or solidarity may regard such approaches as inequitable if they risk exclusion or moral stigma. The framework’s key contribution is to make these normative trade-offs explicit, providing a transparent, evidence-informed structure for deliberation.

Debates around personal responsibility in health policy have largely been conducted at the level of abstract principle, with proponents citing deterrence and fairness and critics pointing to the practical difficulty of distinguishing choice from circumstance (Sharkey et al., 2010, Friesen, 2018, Minkler, 1999). This study moves the debate into the empirical domain. The demonstration that non-vaccination due to personal choice can be systematically distinguished from non-vaccination due to constrained circumstance addresses the objection that choice and circumstance cannot be disentangled in practice in responsibility-based policies (Sharkey et al., 2010, Friesen, 2018).

The study also contributes to approaches that seek a middle ground between holding individuals fully accountable for all health outcomes and disregarding personal choice entirely, such as luck egalitarianism (Segall, 2010, Buyx, 2008, Albertsen et al., 2014). By operationalising Roemer’s distinction between circumstances and effort, and by quantifying the financial consequences in an insured population, this research demonstrates that these principles can be applied in real-world health financing contexts.

While jurisdictions such as Singapore and Quebec invoked the principle that the unvaccinated should bear some of the resulting costs, neither provided an empirical method for distinguishing choice from circumstance nor quantified the excess costs attributable to non-vaccination. This study provides such a method, while recognising that findings from an insured population with comprehensive coverage may not transfer directly to settings where access barriers are more pronounced.

For private insurers, the findings suggest that responsibility-sensitive benefit design can be introduced cautiously and proportionately. Incentive-based mechanisms offer ethically defensible and administratively feasible approaches to promote responsible behaviour while maintaining trust and the integrity of pooled risk. Punitive mechanisms, by contrast, risk undermining both fairness and system cohesion through perceived discrimination, adverse selection, and erosion of member confidence.

For policymakers, particularly for those involved in South Africa’s National Health Insurance (NHI) reforms, these results offer broader lessons. The insured population provides a controlled environment to test responsibility-based approaches but extending them to public systems requires careful consideration of structural inequities and access barriers. The framework offers a practical tool to distinguish between contexts where responsibility is ethically appropriate (where choice is genuine) and contexts where it is not (where opportunities remain constrained).

Debates about personal responsibility reflect a wider dilemma in advancing Universal Health Coverage (UHC) globally: balancing collective solidarity with individual accountability (Minkler, 1999). The COVID-19 experience highlights both the necessity of shared responsibility and the limits of unconditional solidarity when individual behaviour imposes collective costs. This study suggests that responsibility-sensitive justice can complement UHC when applied proportionately and transparently. Framed not as punishment but as cooperation, personal responsibility can encourage individuals to act in ways that sustain equitable and efficient systems. By combining ethical reasoning with empirical data, this research contributes to the emerging discourse on how fairness and efficiency can be integrated rather than opposed, offering a practical means to align autonomy with collective welfare.

### Limitations

Several limitations should be noted. First, the analysis is limited to a privately insured population with relatively homogeneous access to vaccination; findings may not generalise to populations with greater socioeconomic diversity or access barriers. Second, while the decision framework integrates ethical and empirical reasoning, it necessarily simplifies complex moral judgments. Incorporating qualitative insights, such as members’ perceptions of fairness and responsibility, would have strengthened contextual interpretation.

Third, because the synthesis draws on secondary analyses, it also inherits the limitations of each empirical analysis. Observational data are subject to residual confounding, and vaccination status may be misclassified in claims data. The fairness assessment relied on predictive modelling, which may not capture all unmeasured determinants of choice or constraint; in particular, the cost estimates may be affected by unobserved differences in health-seeking behaviour between vaccinated and unvaccinated members, and the binary vaccination classification does not capture the timing of vaccination relative to COVID-19 waves.

Fourth, while the decision framework illustrates how normative reasoning can be structured and applied, it remains conceptual rather than probabilistic; it clarifies how judgments depend on underlying values but does not quantify uncertainty.

Despite these limitations, synthesising evidence across empirical, ethical, and practical domains enhances robustness and validity. The convergence of findings from multiple methodological approaches provides consistent support for the study’s central conclusions, strengthening confidence in the broader inferences dawn.

### Further research

Future research could extend this framework to other contexts where personal responsibility is debated, such as lifestyle-related conditions (e.g., smoking, obesity) or preventive screening behaviours. Applying the framework in public-sector settings would test its relevance under conditions of greater socioeconomic inequality and more constrained access, helping to distinguish contexts where responsibility-sensitive approaches are ethically appropriate from those where they risk reinforcing inequity.

Comparative studies across settings could also assess the framework’s transferability and explore how cultural, institutional, or policy factors shape judgments about fairness and responsibility. Methodologically, combining quantitative and qualitative evidence, stakeholders’ perceptions of fairness, their trust in the system, and their views on procedural legitimacy, would provide a deeper understanding of the public acceptability and ethical reasoning underlying responsibility-based policy decisions.

Finally, further work should explore procedural safeguards to ensure that responsibility-sensitive policy instruments are implemented transparently, proportionately, and in ways that strengthen, rather than weaken, solidarity and trust. As South Africa and other LMICs advance toward Universal Health Coverage, the ability to balance collective protection with individual accountability will remain central to building just and sustainable health systems.

## CONCLUSION

This study applied a structured assessment framework integrating empirical evidence, ethical reasoning, and practical feasibility assessment to evaluate the case for applying personal responsibility for COVID-19 vaccination in a South African insured population. The findings demonstrate that vaccination substantially reduced hospitalisation, utilisation, and expenditure; and that it dominated non-vaccination from a cost-utility perspective. The analysis further showed that non-vaccination due to personal choice can be systematically distinguished from non-vaccination due to constrained circumstance, and that the resulting excess costs can be quantified. Finally, it demonstrated that applying personal responsibility is practically feasible if the approach adopted is proportionate and incremental, starting with low-risk, incentive-based mechanisms.

However, the justification for applying personal responsibility is conditional, context-dependent, and value-sensitive. Prioritising efficiency strengthens the case for responsibility-based mechanisms, while prioritising equity or solidarity weakens it. The legitimacy of responsibility-based measures depends on how fairness, efficiency, and solidarity are prioritised in policy reasoning. Responsibility-sensitive measures are ethically defensible only when applied in settings where choice is demonstrable, benefits are clear, and mechanisms are proportionate, fair, and feasible. Incentive-based and enabling instruments such as premium discounts or wellness rewards emerge as lower-risk starting points than punitive or exclusionary designs which carry higher risks of eroding fairness, equity, and trust.

Beyond COVID-19, these findings offer practical insights for advancing Universal Health Coverage. By showing how personal responsibility can be evaluated and applied within an ethically structured and transparent framework, this research contributes to a pragmatic approach for balancing individual accountability with collective solidarity—foundations essential to fair, efficient, and resilient health systems.

## Declaration of generative AI and AI and AI assisted technologies in the manuscript preparation process

During the preparation of this work the author(s) used ChatGPT and DeepSeek order to assist with language editing, phrasing refinement, and structural clarity. After using this tool/service, the author(s) reviewed and edited the content as needed and take(s) full responsibility for the content of the published article.

## Ethical approval

Ethics approval was granted by the Faculty of Health Sciences Human Research Ethics Committee of the University of Cape Town – approval number HREC Ref: 606/2023.

## Author contributions

Geetesh Solanki, Susan Cleary and Francesca Little designed the study. Geetesh Solanki prepared and ran the analytical models drawn on in this paper with input from Susan Cleary and Francesca Little. Susan Cleary provided critical review oversight. Geetesh Solanki wrote the first draft of this manuscript which was then critically reviewed and revised by the other authors.

## Data Availability

De identified data supporting the findings of this study are available on ZivaHub, the University of Cape Town institutional research data repository. The dataset includes anonymised demographic, vaccination, and claims expenditure records used in the analyses. Data are shared under a Creative Commons Attribution license and may be reused for non-commercial research with appropriate citation.

## Acknowledgements

The authors would like to acknowledge the institutional support to the study authors provided by the University of Cape Town. We also acknowledge NMG Consultant and Actuaries for assisting in providing access to the data and Priya Makanjee for assistance in extracting the data.

